# Aspirin prescribing in cardiovascular disease in middle-aged and older adults in Ireland: findings from The Irish Longitudinal Study on Ageing

**DOI:** 10.1101/2020.07.24.20161703

**Authors:** Frank Moriarty, Alan Barry, Rose Anne Kenny, Tom Fahey

## Abstract

**Background:** Aspirin use for cardiovascular indications is widespread despite evidence not supporting use in patients without cardiovascular disease (CVD). This study characterises aspirin prescribing among people aged ≥50 years in Ireland for primary and secondary prevention, and factors associated with prescription.

**Methods:** This cross-sectional study includes participants from wave 3 (2014-2015) of The Irish Longitudinal Study on Ageing. We identified participants reporting use of prescribed aspirin, other antiplatelets/anticoagulants, and doctor-diagnosed CVD (MI, angina, stroke, TIA) and other cardiovascular conditions. We examined factors associated with aspirin use for primary and secondary prevention in multivariate regression. For a subset, we also examined 10-year cardiovascular risk (using the Framingham general risk score) as a predictor of aspirin use.

**Results:** Among 6,618 participants, the mean age was 66.9 years (SD 9.4) and 55.6% (3,679) were female. Prescribed aspirin was reported by 1,432 participants (21.6%), and 77.6% of aspirin users had no previous CVD. Among participants with previous CVD, 17% were not prescribed aspirin/another antithrombotic. This equates to 201,000 older adults nationally using aspirin for primary prevention, and 16,000 with previous CVD not prescribed an antithrombotic. Among those without CVD, older age, male sex, free health care, and more GP visits were associated with aspirin prescribing. Cardiovascular risk was significantly associated with aspirin use (adjusted relative risk 1.15, 95%CI 1.08-1.23, per 1% increase in cardiovascular risk).

**Conclusion:** Almost four-fifths of people aged ≥50 years on aspirin have no previous CVD, equivalent to 201,000 adults nationally, however prescribing appears rational in targeting higher cardiovascular risk patients.

## Background

Aspirin is one of the most widely prescribed medicines in its role in the management of cardiovascular disease.[1] Due to its inhibitory effects on platelet aggregation and thrombus formation, it is used in the context of secondary prevention (i.e. to prevent further cardiovascular events among patients who have existing cardiovascular disease) and primary prevention (i.e. to prevent the development of cardiovascular disease or a first cardiovascular event).[2] It has also received attention as a potential preventive therapy for cancer, in particular colorectal cancer.[3]

However, evidence on the appropriate cardiovascular indications for prescribing aspirin is equivocal, while evidence on potential cancer prevention benefits is unclear.[4, 5] As further evidence has emerged in relation to aspirin’s benefits and harms, clinical guidelines have varied over time in their recommendations on appropriate indications for prescribing aspirin for cardiovascular treatment and prevention.[2] Up to 2015, treatment guidelines have recommended aspirin be considered for primary prevention among certain higher risk groups. However, since then, guidelines have narrowed the group of patients where use is recommended, or recommended against use for primary prevention. For example, the 2016 European guidelines on cardiovascular prevention recommend against antiplatelet agents in patients without cardiovascular disease history (primary prevention).[6] The US Preventive Services Task Force 2016 recommendations to initiate aspirin in adults aged 50 to 59 years with a ten year cardiovascular disease risk of 10% or higher who have no increased bleeding risk and a life expectancy of at least 10 years.[7] For those aged 60-69 years, people meeting the above criteria are more likely to benefit, but the prescribing decision ought to be made at the individual level. These recognise the need to balance potential benefits against known adverse effects, particularly bleeding risks.

Much of the evidence was derived from trials conducted before the year 2000, however, three large randomised trials were published in 2018 evaluating aspirin in primary cardiovascular prevention[8–10]. Considering the evidence from the “modern era” suggests that while the harms of aspirin with respect to bleeding risks have remained stable, the absolute benefits have reduced over time (potentially due to decreased baseline cardiovascular risk), and use for primary prevention should be reconsidered.[11] Despite this, aspirin is widely used, particularly in older age groups who may have increased cardiovascular risk but also increased risk of haemorrhagic adverse effects. Use where not indicated, as well as omission among people with a previous cardiovascular event, have been flagged as potentially inappropriate prescribing in STOPP/START.[12] Aspirin overprescribing may have significant implications at the population level in terms of medication harms and costs. This may be compounded by lack of discontinuation among patients no longer in a recommended group, either due to change in guidelines or individual ageing and changing balance of benefits and risks.

In many health systems, low-dose aspirin for cardiovascular prevention is available without a prescription as an over-the-counter medication. However, in Ireland, it is not available over-the-counter and must be prescribed by a doctor. Therefore, this study aims to characterise prescribing of aspirin among people aged ≥50 years in Ireland for primary and secondary prevention. The objectives are to determine:

1. Prevalence of aspirin use for primary and secondary prevention.
2. Factors associated with prescription of aspirin in those with and without previous cardiovascular disease.
3. Whether cardiovascular risk is associated with aspirin prescribing for primary prevention.

## Methods

This is a cross-sectional study of respondents to wave 3 (2014-2015) of The Irish Longitudinal Study on Ageing (TILDA). TILDA is a nationally-representative cohort study examining the health, economic and social circumstances of middle-aged and older people living in the community.[13] Ireland has a mixed public-private health system. A proportion of the population are eligible for a range free at the point-of-care health services, based on household income and age. Those not covered under the general practitioner (GP) visit or medical card schemes are considered private patients and pay for GP visits and other health services. TILDA recruited approximately 8,000 people in the Republic of Ireland, based on a random selection process using a national geodirectory of residential addresses. Baseline recruitment and data collection occurred between 2009-2011 and involved a face-to-face computer assisted personal interview (CAPI) conducted by a trained interviewer in the respondent’s home, a self-completion questionnaire, and a health assessment conducted either at home or at a test centre on a subset of respondents. Follow-up of participants is completed every two years, with the health assessment repeated every four years (alternate waves).

Information on medication use is gathered in TILDA during the CAPI by asking respondents to show all medications which they take on a regular basis, such as every day or week. Medication brand names are recorded and these entries are linked to the corresponding World Health Organisation Anatomical Therapeutic Classification (ATC) code and non-proprietary or drug name. Aspirin prescribing will be identified from self-reported medications using the ATC codes ‘B01AC06’ and ‘N02BA01’ and instances where an over-the-counter brand of aspirin (indicated for analgesia) is specified will be excluded. Low-dose aspirin (the formulation licensed for cardiovascular prevention and treatment) is not available over-the-counter in Ireland.

Other variables included in the present study are age, sex, educational attainment (as a proxy of socioeconomic status), area of residence, health cover, reported number of GP visits in the previous 12 months, and specific cardiovascular conditions. We considered Dublin separately from other urban areas due to the higher density of hospitals here. Respondents are asked if a doctor has ever told them they have various named cardiovascular and other health conditions, including myocardial infarction (MI), stroke, hypertension, and diabetes. Following this, respondents are asked if they have any other cardiovascular conditions and these free-text responses were screened to identify further cardiovascular conditions. Dates of cardiovascular events are not captured, and therefore appropriateness of dual antiplatelet therapy could not be assessed. As well as considering these morbidities individually, respondents were classified as eligible for secondary prevention if they report previous cardiovascular disease (i.e. MI, angina, coronary artery disease or cerebrovascular disease (stroke and transient ischemic attack) or as eligible for primary prevention if they report no previous cardiovascular disease. For a sub-group of participants aged between 50 and 74 years who underwent an objective health assessment, we calculated their predicted 10-year cardiovascular risk based on the Framingham General Cardiovascular Risk score for use in primary care.[14] This estimates an individual’s risk of all potential manifestations and adverse consequences of atherosclerosis, including fatal and non-fatal events, and was calculated using age, gender, smoking status, low-density lipoprotein cholesterol, and blood pressure (based on the mean of two measurements taken one minute apart while the participant was seated).

### Analysis

Characteristics of included participants were described overall, and separately for those who reporting being prescribed aspirin and those who did not. Use of aspirin and other antithrombotics was summarised across individual cardiovascular conditions. This was also examined across categories when participants were assigned to the most serious cardiovascular condition they reported, based on a previously developed indication hierarchy.[15] Lastly, use was summarised for those eligible for primary and secondary prevention. Population weights, based on age, sex, highest level of education attained and urban/rural residence distribution of the population of Ireland in the 2011 Census, were applied to estimate the number of individuals nationally with no previous cardiovascular disease receiving aspirin for primary prevention, and those with previous cardiovascular disease not receiving anti-thrombotic therapy.

Then, multivariate generalised linear regression models were used to determine factors associated with aspirin use (binary) in the primary prevention cohort, and aspirin or other antithrombotic use in the secondary prevention cohort. Covariates included age, sex, education, location, health cover status, and frequency of GP visits (divided into quintiles). An age group and sex interaction was evaluated to estimate how probability of aspirin/antithrombotic use varied.

Last, among respondents who completed a health assessment and had no previous cardiovascular disease (i.e. primary prevention cohort), their calculated cardiovascular risk was compared between aspirin users and non-users. This was then included in the multivariate regression model to assess its relationship with aspirin use, adjusting for other patient factors.

## Results

### Descriptive characteristics

A total of 6,618 participants from wave 3 of TILDA were included in this study (see Table 1), of whom 55.6% (3,679) were female with a mean age of 66.9 years (SD 9.4). Slightly more than half (52.2%) were eligible for a medical card or GP visit card, and the median number of GP visits was 3 (interquartile range 2-5).

**Table 1.** Characteristics of all included participants, and separately for those who did and did not report prescribed aspirin.

| Characteristics | n (%) |  |  |
| --- | --- | --- | --- |
|  | No aspirin (n=5186) | Aspirin (n=1432) | Total (n=6618) |
| **Age** |  |  |  |
| Mean (SD) | 65.6 (9.1) | 71.6 (9.0) | 66.9 (9.4) |
| 50-64 years | 2702 (52.1) | 334 (23.3) | 3036 (45.9) |
| 65-74 years | 1556 (30.0) | 554 (38.7) | 2110 (31.9) |
| ≥75 years | 928 (17.9) | 544 (38.0) | 1472 (22.2) |
| **Female sex** | 3049 (58.8) | 630 (44.0) | 3679 (55.6) |
| **Education** |  |  |  |
| Primary/none | 1208 (23.3) | 529 (37.0) | 1737 (26.3) |
| Secondary | 2098 (40.5) | 512 (35.8) | 2610 (39.4) |
| Third/higher | 1879 (36.2) | 390 (27.3) | 2269 (34.3) |
| **Area of residence** |  |  |  |
| Dublin city or county | 1241 (23.9) | 351 (24.5) | 1592 (24.1) |
| Another town or city | 1410 (27.2) | 430 (30.0) | 1840 (27.8) |
| A rural area | 2535 (48.9) | 651 (45.5) | 3186 (48.1) |
| **GP Visit or medical card holder** | 2432 (47.0) | 1020 (71.3) | 3452 (52.2) |
| **Annual GP visits** |  |  |  |
| Median (IQR) | 2 (1, 4) | 4 (2, 6) | 3 (2, 5) |
| 1st quintile | 1469 (28.4) | 163 (11.4) | 1632 (24.7) |
| 2nd quintile | 1180 (22.8) | 275 (19.3) | 1455 (22.1) |
| 3rd quintile | 1336 (25.9) | 507 (35.6) | 1843 (27.9) |
| 4th quintile | 252 (4.9) | 98 (6.9) | 350 (5.3) |
| 5th quintile | 931 (18.0) | 383 (26.9) | 1314 (19.9) |

Prescribed aspirin was reported by 1,432 participants (21.6%). Considering cardiovascular conditions individually, the highest prevalence of use was in those with a previous MI (72.8%), and when participants were classified according to the most serious cardiovascular indication they reported, the prevalence decreased moving down the indication hierarchy (Figure 1A). Among those with previous cardiovascular disease (previous MI, angina, stroke, or TIA), 16.5% were not prescribed aspirin or another antithrombotic. The prevalence of reported gastric ulcer history did not differ significantly between those prescribed and not prescribed an antithrombotic for secondary prevention. In absolute terms, the majority of aspirin users were in the lower levels of the indication hierarchy (Figure 1B). In 77.6% of cases, aspirin was prescribed for primary prevention i.e. to people with no previous cardiovascular disease (Figure 1C). Weighting the TILDA cohort to the national population, these estimates equate to 16,000 middle-aged and older adults nationally with previous cardiovascular disease not prescribed aspirin or another antithrombotic, and 201,000 people prescribed aspirin for primary prevention.

**Figure 1.**
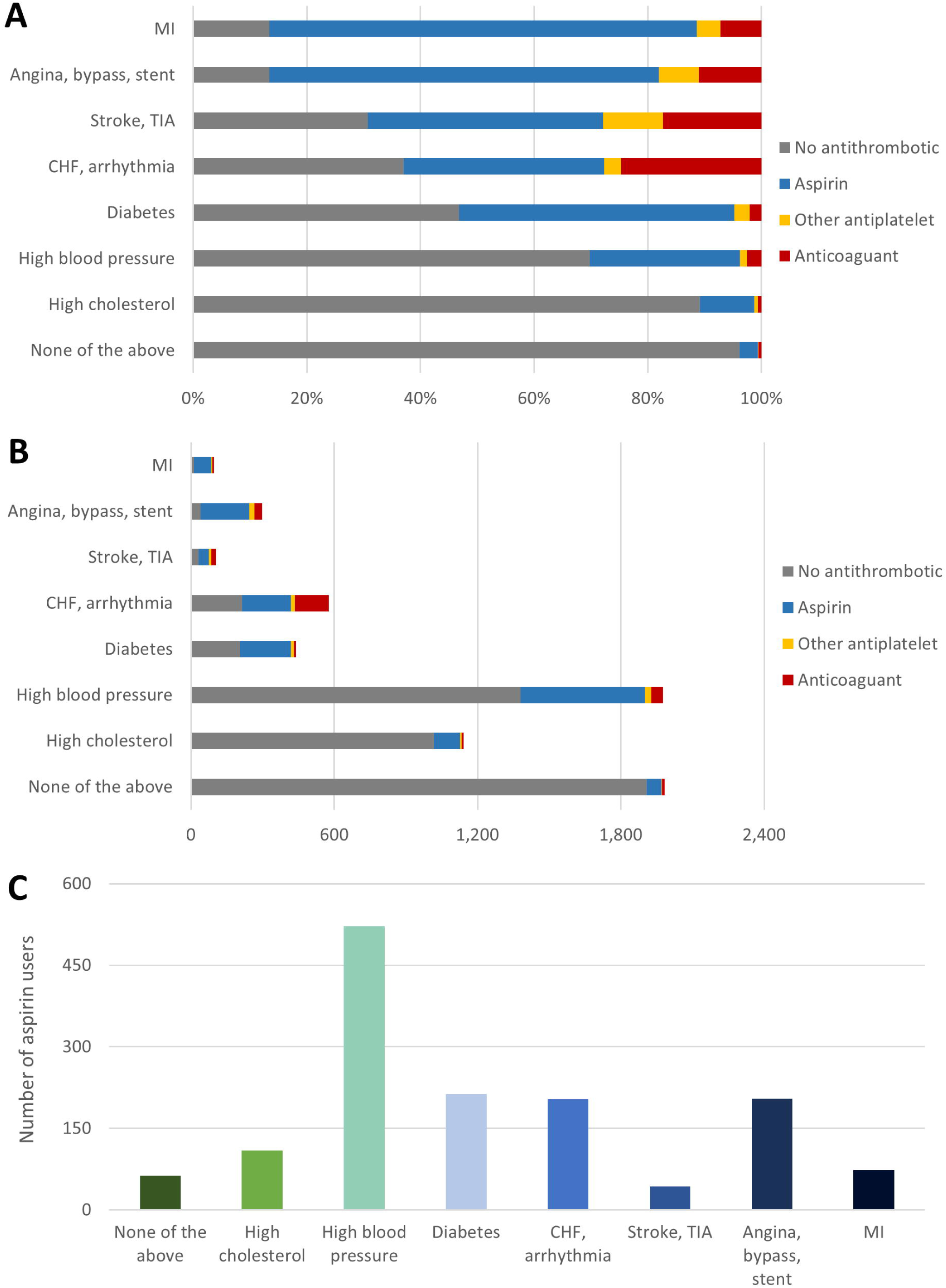
Distribution of aspirin and other antithrombotic use across indication hierarchy categories, in percentage (A) and absolute (B) terms; distribution of aspirin users across indication hierarchy categories (C). CHF – congestive heart failure; MI – myocardial infarction; TIA – transient ischaemic attack.

### Factors associated with aspirin prescribing

Among the primary prevention cohort, older age, male sex, having a GP visit or medical card, and more GP visits were associated with aspirin prescribing (Figure 2). When an age group-sex interaction was considered, the difference in the likelihood of aspirin prescription between men and women was less in higher age groups (eFigure 1). For the secondary prevention cohort, use of aspirin or another antithrombotic was associated with male sex (Figure 2), and there was no significant age group-sex interaction (eFigure 2).

**Figure 2.**
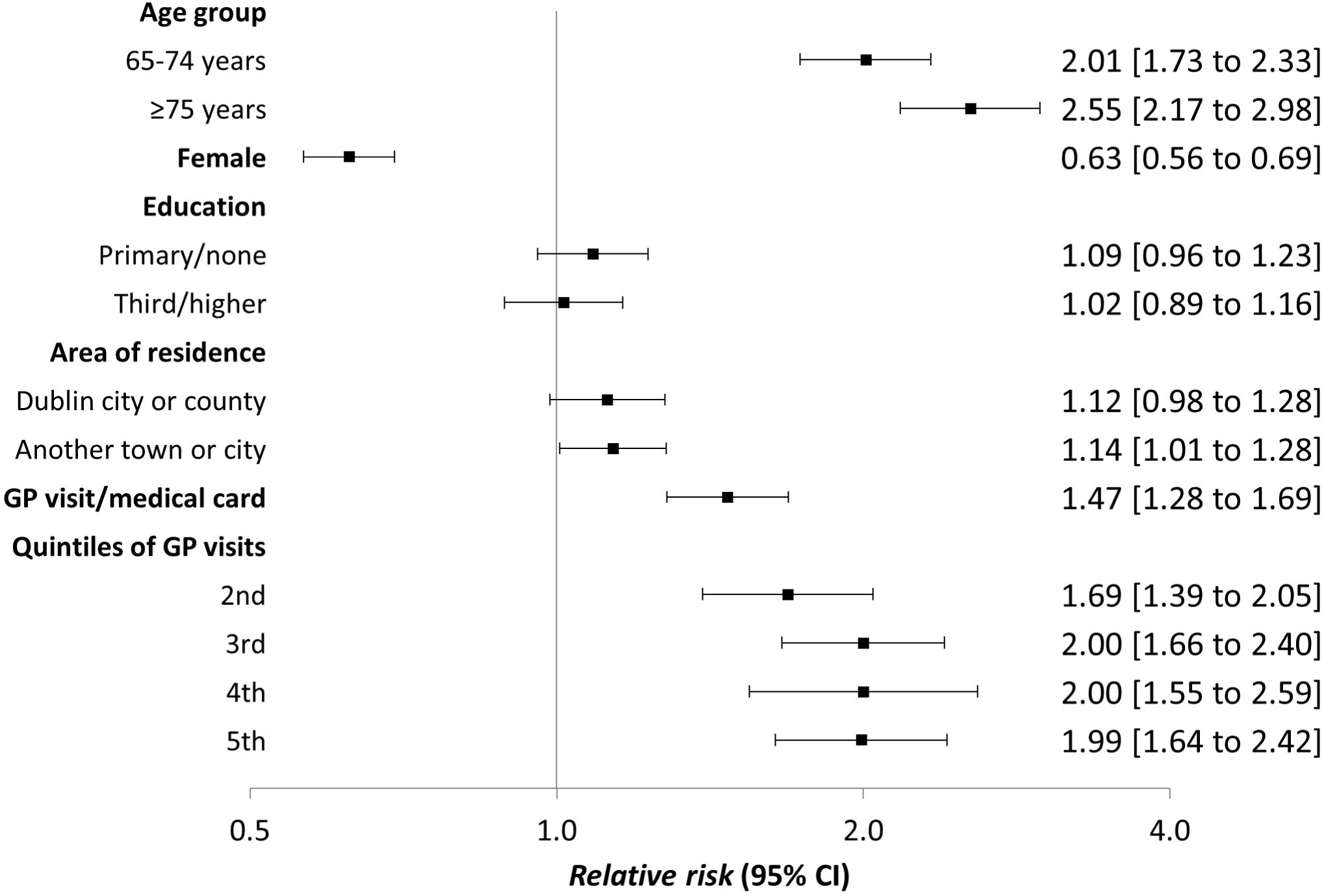
Factors associated with aspirin prescription among participants with no previous cardiovascular events (primary prevention) (n=6,091)

**Figure 3.**
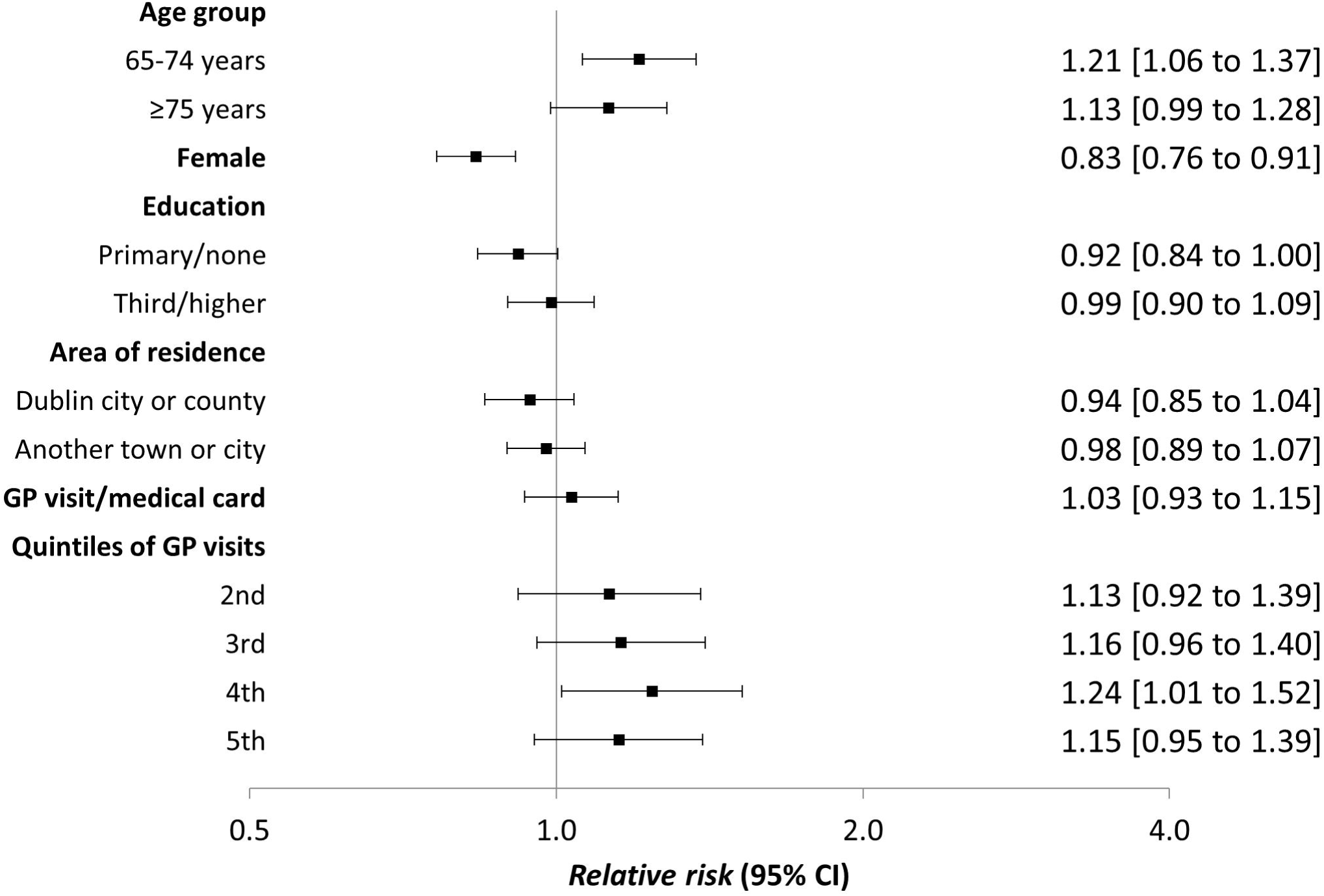
Factors associated with aspirin or another antithrombotic prescription among participants with previous cardiovascular disease (secondary prevention) (n=494)

### Framingham general risk assessment

Among the 3,784 participants with no previous cardiovascular disease who underwent a health assessment, the mean predicted cardiovascular risk was higher amongst those prescribed aspirin (21.4%) compared to non-users (14.6%). Cardiovascular risk remained a significant predictor of aspirin use among the primary prevention cohort after adjustment for demographic characteristics, with a 1% increase in Framingham General Cardiovascular Risk being associated with a 15% (95%CI 8 to 23%) increased likelihood of aspirin prescription (Table 2).

**Table 2.** Association of Framingham General Cardiovascular Risk with aspirin prescription among participants with no previous cardiovascular disease who underwent a health assessment (n=3,784)

|  | Relative risk of aspirin prescription* (95% CI) |
| --- | --- |
| **Framingham risk (per 1% change in risk)** | 1.15 (1.08 to 1.23) |
| **Framingham risk categories** |  |
| < 10% | 1.00 (ref) |
| 10% to 19.9% | 1.87 (1.46 to 2.40) |
| ≥ 20% | 2.32 (1.72 to 3.14) |

\*adjusted for age, sex, education, area of residence, health cover, GP utilisation

## Discussion

We found that in this middle-aged and older cohort of community dwellers in Ireland, aspirin was prescribed for primary prevention in a large majority of cases, where evidence suggests the benefit/harm ratio is unfavourable. This prescribing was directed at patients with higher cardiovascular risk. Considering individuals with previous cardiovascular disease where there is a clear indication for secondary prevention, almost 1 in 5 were not being treated with aspirin or another antithrombotic agent.

### Comparison with the literature

A number of individual characteristics were associated with aspirin prescription among those with no previous cardiovascular disease, including entitlement to free GP care and higher frequency of GP visits. Entitlement to free healthcare can contribute to increased frequency of visits,[16] and thus more opportunities for a doctor to prescribe, as has been show previously for prescribing of statins,[17, 18] and aspirin.[19] A US survey found physicians with higher public insurance (Medicaid) income were more likely to recommend prescribing of statins for primary prevention, a scenario where they were not recommend by contemporary guidelines.[20] In an Irish context, free healthcare was shown to be associated with polypharmacy,[21] and while this relationship may be subject to confounding by socioeconomic status and other factors, the association remained after multivariate adjustment or when propensity score matching was used. As Ireland is currently moving toward a system of universal entitlement to primary care free at the point of access under Sláintecare,[22] it is important to consider how this may have implications for effective use of care and potential for overprescribing.

Although its use is not supported by evidence and not recommended by the latest European Society of Cardiology guidelines,[6] prescribing for primary prevention in this study appeared to be rational in targeting those with higher cardiovascular risk. Similar has been found in studies in the US (for example, where a survey of adults found 40.9% of those with high cardiovascular risk were aspirin by their doctor, compared to 26.0% in the lower risk group),[23, 24] as well as in Italian and Swiss setting.[25, 26] However, in common with the present study, these studies found there was still substantial use for primary prevention among those with low cardiovascular risk. A previous analysis of STOPP/START criteria in earlier waves of TILDA, based on self-reported diagnoses and administrative medication dispensing data, identified the criteria relating to prescribing in primary prevention in 19.6% of participants aged ≥65 years, while omission in secondary prevention had a prevalence of 2.6%.[27] Consistent with previous literature, we found women with previous CVD were less likely to be on an antithrombotic,[25] reinforcing the need to address the well-established underdiagnosis and undertreament of CVD in women. Although it was not possible to examine in our study, racial disparities in aspirin use have also previously been shown.[28, 29]

### Implications

Extensive use of aspirin for primary prevention (in 80% of people on aspirin aged 50 years and up) has implications both for costs (to patients and the health system) and patient outcomes. The health system in Ireland paid approximately €20 million for just over 3 million aspirin dispensings on state drug schemes in 2017, with further out-of-pocket expenditure likely among those not covered by these schemes.[30] In terms of patient outcomes, evidence suggests low absolute benefits associated with aspirin use compared to the absolute harms. For 1,200 individuals taking aspirin for primary prevention for five years, there would be four fewer major adverse cardiovascular events or MACEs (cardiovascular death, or non-fatal stroke or MI) and three fewer ischaemic strokes, but also three more intracranial haemorrhages and eight more major bleeding events.[11] Applying these estimates to the 201,000 users for primary prevention in the present study, this equates to 670 MACEs averted over five years, at a cost of approximately 500 intracranial haemorrhages and 1,340 major bleeding events in Ireland’s middle-aged and older population per year. This is substantially higher than the 700 gastric bleeds and haemorrhagic strokes over 10 years attributable to aspirin overprescribing in a Swiss population study.[26]

A patient’s decision to start or continue taking aspirin, while potentially influenced by a doctor’s recommendation,[31] should be informed by knowledge of the benefits and risks. These can substantially affect willingness to take a medicine. A study of older adults found that for a hypothetical medication that reduced myocardial infarction risk by six events per 100 people, 48-69% were unwilling/uncertain about taking if it cause daily mild adverse effects (fatigue, dizziness, nausea, slowed thinking).[32] This increased to 88% for adverse effects that affected functioning. The risk of serious bleeding events should therefore be discussed with patients.

The high level of aspirin use for primary prevention may partly represent a legacy of historical guideline recommendations,[2] which could be investigated in future research using prescriber vignettes. For instance, our finding of high numbers of aspirin users with hypertension may have been influenced by the Hypertension Optimal Treatment trial.[33] The scenario where guidelines have changed and patients on aspirin are no longer recommended to take it presents a challenge for these patients, and their doctors, in deciding whether to continue or stop aspirin in the face of new evidence. Similar is true for patients who remain on aspirin for primary prevention over time and reach an age where it is not recommended. Observational studies of aspirin discontinuation among patients without cardiovascular disease have been conflicting on whether this increases risk of cardiovascular events.[34, 35] However, studies using routine data may be unable to differentiate between deprescribing (i.e. a clinician supervised reducing or stopping where it is judged that risks outweigh benefits) and discontinuation due to adverse events or poor adherence, and they may also be subject to residual confounding. To date, it appears no interventional studies of aspirin deprescribing in primary prevention have been conducted. A deprescribing guideline for aspirin has been developed to support patients and prescribers in assessing the need for ongoing treatment, and recommending optimal approaches for deprescribing,[36] however, such deprescribing recommendations could also be incorporated as part of standard clinical guidelines.[37]

### Strengths and limitations

We used the Framingham General Cardiovascular risk score as it is suitable for use in adults up to 74 years of age, however, there is evidence of poor calibration (where it overestimates risk), but its discrimination is reasonable (i.e. ability to differentiate those at different levels of risk) which is most relevant for its use in this study.[38] A strength is participants can report use of aspirin regardless of whether it is prescribed or not. Although low dose aspirin for cardiovascular prevention is prescription only in Ireland, cases where an individual is purchasing over-the-counter medication from other jurisdictions can be captured as opposed to using only administrative prescribing or dispensing data which may result in misclassification. Self-report of both medication use and cardiovascular morbidity may introduce bias, however, previous validation has shown good agreement in TILDA between reported and dispensed medications.[39] We would expect high participant recall of major cardiovascular events, such as myocardial infraction and stroke,[40] and therefore underreporting of previous cardiovascular disease and misclassification as eligible for primary prevention to be low. Lastly the generalisability of the findings may be limited to Ireland, however, the TILDA cohort are nationally representative.

## Conclusions

Aspirin is commonly prescribed among middle and older-aged adults in Ireland, however, there is still potential to optimise use of it and other antithrombotics in those with existing cardiovascular disease. Most aspirin prescriptions are for those without previous cardiovascular disease. Although prescribing is targeted at those with higher cardiovascular risk, prescribers should reconsider starting aspirin for primary prevention for patients where risks are likely to outweigh benefits, and whether current users should continue on aspirin.

## Data Availability

Data from TILDA can be requested via https://tilda.tcd.ie/data/accessing-data/

## Summary box

What is already known on this subject?

- Evidence from recent randomised controlled trials suggests low-dose aspirin does not provide a net benefit in people with no previous cardiovascular disease (i.e. primary prevention).
- Aspirin is one of the most commonly used medicines, however it can be unclear what indication it is being prescribed for and factors that affect its use.

What does this study add?

- In Ireland, high proportions of middle-aged and older people with previous cardiovascular disease are prescribed aspirin or an antithrombotic, but equivalent to 16,000 people nationally are not treated.
- The majority of aspirin prescribing appears to be for primary prevention, equivalent to 201,000 people nationally.
- Although predicted cardiovascular risk was associated with aspirin prescribing for primary prevention, risks and benefits should be weighed up when considering aspirin for cardiovascular prevention.

## Acknowledgements

We wish to acknowledge all those who participated in The Irish Longitudinal Study on Ageing.

## Contributors

FM conceived the study, and all authors were involved in study design. RAK acquired the study data, FM and AB completed analysis and all authors interpreted the data. FM prepared the initial draft, and all authors were involved in the critical revision of this and approval of the final manuscript

## Ethics approval

Trinity College Dublin Faculty of Health Sciences Research Ethics Committee.

